# Declines in Influenza Vaccination Coverage in White and Black, Non-Hispanic Children from 2012-2019 to 2019-2022

**DOI:** 10.1101/2023.06.08.23290565

**Authors:** Abigail L. Corle

## Abstract

Declines in routine influenza vaccination rates have become a cause for concern as influenza vaccine coverage rates have declined among Black, Non-Hispanic children compared to an increase in coverage among White, Non-Hispanic children ages 6 months to 17 years old. Influenza season years analyzed were 2012-2022 using data provided by the US Center for Disease Control’s FluVaxView database. Data for this database is sourced from the National Immunization Survey-Flu (NIS-Flu) and the Behavioral Risk Factor Surveillance System (BRFSS) survey. White, Non-Hispanic children saw an increase in vaccination coverage from 55.8% to 60.3%, whereas Black, Non-Hispanic children decreased from 58.2% to 52.9% coverage. Chi-squared tests with Yates correlation were performed to find statistical significance. Vaccination coverage rate changes for both demographics were found to be statistically significant at p < 0.00001. 70% influenza vaccination coverage rate remains the US Department of Health’s *HealthyPeople 2030* initiative’s goal for all demographics and may be achieved by utilizing interventional tools such as motivational interviewing, provider-patient dialogue, community-based vaccination clinics, or home visits. Evidence-based interventional provider-patient dialogue has had success in increasing vaccination rates among other vaccines. Continued research into the cause of influenza vaccination rate decline among vulnerable and medically underserved populations is needed to best implement vaccine-based interventions.

## Introduction

The influenza vaccine is a routine vaccine that has increased in importance in a post-COVID-19 world. During the COVID-19 pandemic, numerous campaigns were held to increase vaccination coverage for the influenza vaccine. Despite this, low vaccination rates of 52.9% coverage have been observed for Black, non-Hispanic children in the 2019-2022 influenza seasons (1). The aforementioned low vaccination rate value is a substantial decrease from the 58.2% on average in the 2012-2019 influenza seasons (1). Concern has been expressed for an influenza rebound following the COVID-19 pandemic that may lead to difficulties in controlling future influenza seasons (2). It is known that Black communities face a problem of health inequity, where vaccinations may be hindered by lack of transportation or information, geographic segregation, or cost as compared to White communities (3). Between 2012-2019 and 2019-2022, influenza vaccination rates rose among White non-Hispanic children, beginning at 55.8% and ending at 60.3% (1). Despite the many workforce hardships caused by COVID-19, it is known that the low vaccination rates were not caused by a lack of influenza vaccine supply (4).

Children ages 6 months to 17 years currently account for the largest amount of outpatient visits regarding respiratory illnesses. Additionally, influenza-related pediatric deaths have reached a 3 year high of 152 as of the week of May 19^th^, 2023 (5). More research is required if this rate of influenza vaccine coverage has decreased enough to warrant additional actions (such as actively promoting vaccines) among the providers that care for Black, non-Hispanic children. This data-analysis was performed in order to investigate the significance of the low influenza vaccination rates among Black, non-Hispanic children as compared to White, non-Hispanic children.

## Methods

Datasets provided by the Center for Disease Controls’ Flu Vaccine Coverage database, FluVaxView, were analyzed. FluVaxView provides yearly data regarding percentage coverage of influenza vaccination in both children and adults based upon specific age, sex, race/ethnicity, geographical location (within the United States), and place of vaccination. FluVaxView compiles its data from National Immunization Survey-Flu (NIS-Flu) and Behavioral Risk Factor Surveillance System (BRFSS) surveys. BRFSS conducts its surveys largely through in-person interviews, whereas the NIS-Flu collects its data via State Inpatient Databases created by the Healthcare Cost and Utilization Project (1,4). The exposure of interest is the impact of the COVID-19 pandemic. Using the “Percentage Vaccinated” variable, I analyzed the CDC FluVaxView database’s change in the average vaccination coverage percentage in the target populations of White and Black, non-Hispanic children ages 6 months to 17 years old within the ranges of 2012-2019 and 2019-2022. To best protect the rights of human subjects, I have completed the required CITI research and ethics training and have created an Institutional Review Board account. The study was determined to be ‘Not Human Subject Research’ by the IRB.

Variation was utilized as a form of descriptive analysis to detect any major loss or gain in percentage vaccination coverage within the target population of both White and Black, non-Hispanic children ages 6 months to 17 years old and depicted using a line series graph. All calculations were performed and compiled using Microsoft Excel 2023. Data was further analyzed using a chi-squared test without Yates correction and used to calculate the p-value. Chi-squared values will be determined using percentages of vaccination coverage given by FluVaxView.

## Results

Influenza vaccination coverage percentage among Black, non-Hispanic children showed a decrease during 2019-2022 influenza seasons (58.2%) compared to pre-pandemic influenza seasons of 2012-2019 (52.9%) in Figure 1; vaccination coverage for White, non-Hispanic children largely increased from 55.8% to 60.3% as shown in Figure 1. Table 1 shows the sample size and percentage vaccinated for each influenza season by year for both demographics. The Black, non-Hispanic chi-squared test in table 2 showed a p-value X^2^(1, N = 137467) = 117.97, p < 0.00001. The White, non-Hispanic chi-squared test in table 3 also showed an identical, p value X^2^(1, N = 795636) = 2337.9, p < 0.00001.

**Table 1.** Compiled sample sizes and percentages of vaccinated children for each influenza season from 2012-2019 and 2019-2022.

|  | White, Non-Hispanic Children |  | Black, Non-Hispanic Children |  |
| --- | --- | --- | --- | --- |
| Year | Percentage Vaccinated | N | Percentage Vaccinated | N |
| 2012-2013 | 53.8 | 61,350 | 56.7 | 10,414 |
| 2013-2014 | 55.2 | 75,934 | 57.2 | 12,934 |
| 2014-2015 | 56 | 74,683 | 58.3 | 13,368 |
| 2015-2016 | 55.3 | 73,660 | 60.9 | 13,717 |
| 2016-2017 | 54.3 | 100,341 | 59.3 | 14,196 |
| 2017-2018 | 55.6 | 69,289 | 55.3 | 11,919 |
| 2018-2019 | 60.5 | 80,684 | 59.5 | 13,171 |
| 2019-2020 | 63.1 | 88,926 | 57.9 | 14,517 |
| 2020-2021 | 60.4 | 82,087 | 49.1 | 15,105 |
| 2021-2022 | 57.4 | 88,682 | 51.6 | 18,126 |

**Table 2.** Results calculated from data provided by NIS-Flu and BRFSS comparing the sample sizes of Black, non-Hispanic children ages 6 months to 17 years. Data is stratified by influenza vaccination status and influenza season years.

|  | Vaccinated | Unvaccinated | p-value |
| --- | --- | --- | --- |
| Pre-Pandemic (2012-2020) | 52216 | 37503 | < 0.00001 |
| Pandemic (2020-2022) | 25259 | 22489 |  |

**Table 3.** Results calculated from data provided by NIS-Flu and BRFSS comparing the sample sizes of White, non-Hispanic children ages 6 months to 17 years. Data is stratified by influenza vaccination status and influenza season years.

|  | Vaccinated | Unvaccinated | p-value |
| --- | --- | --- | --- |
| Pre-Pandemic (2012-2020) | 299055 | 236886 | < 0.00001 |
| Pandemic (2020-2022) | 156596 | 103099 |  |

**Figure 1.**
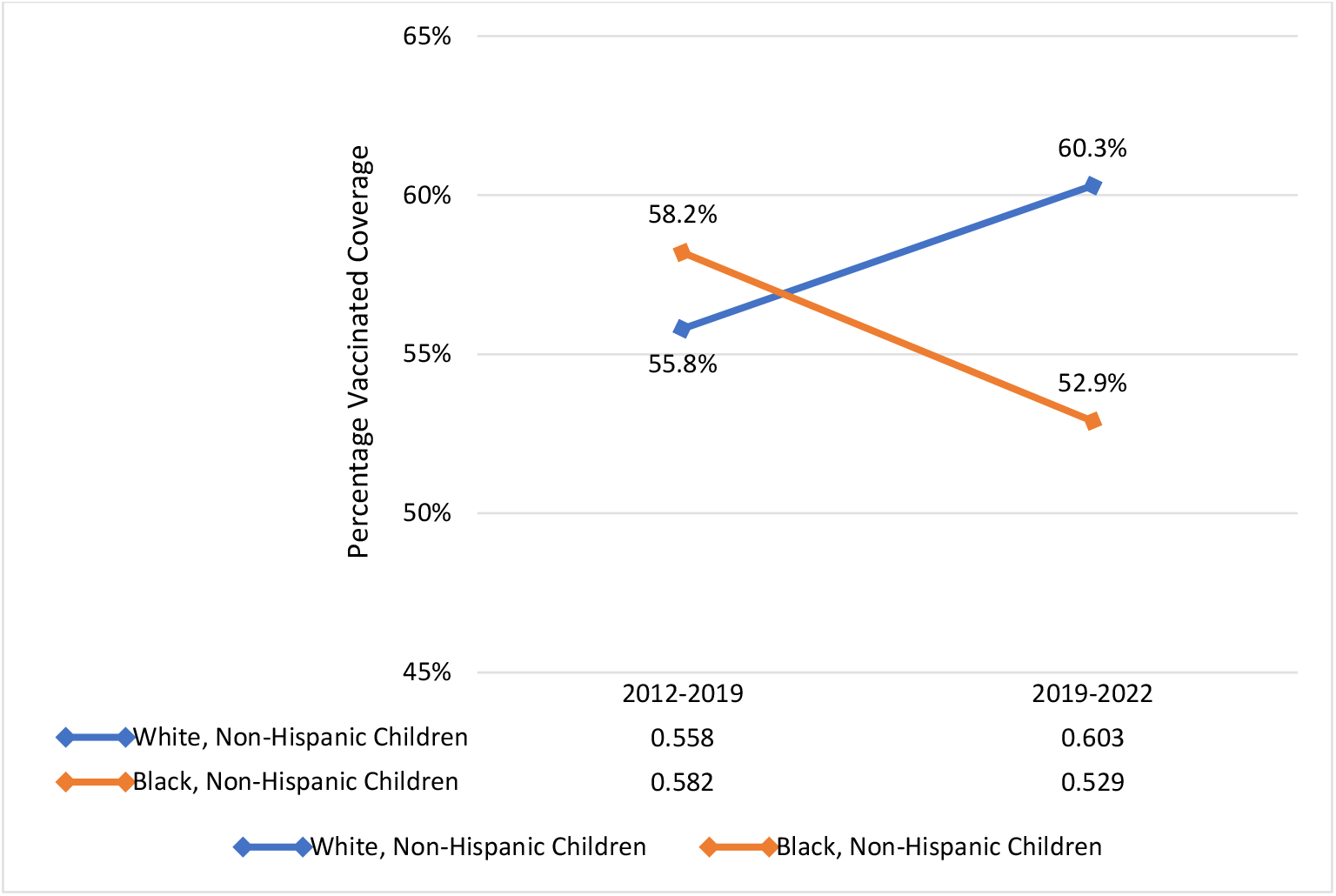
Figure showing the percentage of influenza vaccine coverage in White and Black, non-Hispanic children ages 6 months to 17 years.

## Discussion

A noticeable decrease in influenza vaccination rates was present among Black, Non-Hispanic children following the advent of the COVID-19 pandemic in 2020 compared to their pre-pandemic levels in the 2012-2019 flu seasons. The final vaccination coverage value is far below the United States Department of Health *HealthyPeople 2030* initiative’s desired target of 70% coverage by 2030 among all populations to better protect communities (6). The percentage coverage among White, Non-Hispanic children is also below this threshold, however there is a greatly positive trend towards the goal. By partnering with community-based resources, influenza vaccinations may be more attainable for Black, non-Hispanic communities that experience health inequities that may be absent among White communities (7). With mobile healthcare becoming more common, home visits for vaccination are a method that could increase vaccination coverage if medical clinics are inaccessible (8).

### Clinical Significance

Due the significance of the found p values, it is important to consider more personalized medicine to raise the vaccination rates among both demographics towards the desired target of 70% coverage. Motivational interviewing may be an effective patient-centered tool to convince patients of the benefits of vaccines and to dispel any myths the patient may believe regarding vaccines. Motivational interviewing entails creating an empathy-based partnership between the patient and the provider to foster a healthy learning environment for the patient to learn about vaccines without being judged or scrutinized (9). This style of provider-patient communication has been effective for other vaccines such as many neonatal vaccines, with vaccination coverage increasing as much as 15% across 3, 5, and 7 months of age (10).

## Conclusion

A decrease in vaccinations among all demographics has been cause for concern in the current medical climate. Health inequities among Black, non-Hispanic children have been increasingly prevalent due to the COVID-19 pandemic, leading to decreased average influenza vaccination coverage as compared to White, non-Hispanic children. Inaccessibility to clinics and distrust in modern medicine have contributed to this decrease, showing the importance of community-based healthcare and medical education for those in communities experiencing health inequities to reverse the decrease in influenza vaccination coverage.

Increased attention towards community-based vaccination clinics and home visits may provide an accessible avenue for those with limited medical care availability. More individualized medicine may also prove effective if community-based education is already in place. Evidence-based provider-patient dialogue is an effective tool utilized to increase vaccination coverage. Improving and combining individual-centered care and community-based education approaches is recommended to reach underserved communities and increase vaccination rates.

## Data Availability

All data produced are available on request to the author.

https://www.cdc.gov/flu/excel/fluvaxview/2022/OnlineReport_2021_22_AdditionalTable_FluVxByRaceAndAge_Children.xlsx

https://www.cdc.gov/flu/excel/fluvaxview/2021/general_population/OnlineReport_2020_21_AdditionalTable_FluVxByRaceAndAge_Children-10_5_21.xlsx

https://www.cdc.gov/flu/excel/fluvaxview/2020/general_population/OnlineReport_2019_20_AdditionalTable_FluVxByRaceAndAge_Children-v2.xlsx

https://www.cdc.gov/flu/excel/fluvaxview/2019/general_population/OnlineReport_2018_19_AdditionalTable_FluVxByRaceAndAge_Children-cor-ver.xlsx

https://www.cdc.gov/flu/excel/fluvaxview/2018/children/OnlineReport_2017_18_AdditionalTable_FluVxByRaceAndAge_Children-Cor-Ver.xlsx

https://www.cdc.gov/flu/excel/fluvaxview/2017/general_population/coverage-supplement-table-6.xlsx

https://www.cdc.gov/flu/excel/fluvaxview/2016/2015-16_coverage_supplemental-1.xlsx

https://www.cdc.gov/flu/excel/fluvaxview/sep-2015/2014-15_coverage_supplemental-1.xlsx

https://www.cdc.gov/flu/excel/fluvaxview/sep-2014/2013-14_coverage_supplemental-1.xlsx

https://www.cdc.gov/flu/excel/fluvaxview/2012-13_coverage_supplemental-1.xlsx

## Acknowledgements

I am eternally grateful to Dr. Brian Piper for his feedback, guidance, and tutelage in the creation of this manuscript. Furthermore, I would like to thank the VaxView team with the US CDC for their assistance in acquiring the proper data sets.

